# Cost-benefit of limited isolation and testing in COVID-19 mitigation

**DOI:** 10.1101/2020.04.09.20059790

**Authors:** Andreas Eilersen, Kim Sneppen

## Abstract

The international community has been put in an unprecedented situation by the COVID-19 pandemic. Creating models to describe and quantify alternative mitigation strategies becomes increasingly urgent. In this study, we propose an agent-based model of disease transmission in a society divided into closely connected families, workplaces, and social groups. This allows us to discuss mitigation strategies, including targeted quarantine measures. We find that workplace and more diffuse social contacts are roughly equally important to disease spread, and that an effective lockdown must target both. We examine the cost-benefit of replacing a lockdown with tracing and quarantining contacts of the infected. Quarantine can contribute substantially to mitigation, even if it has short duration and is done within households. When reopening society, testing and quarantining is a strategy that is much cheaper in terms of lost workdays than a long lockdown of workplaces. A targeted quarantine strategy is quite efficient with only 5 days of quarantine, and its effect increases when testing is more widespread.

## Introduction

The 2020 coronavirus (COVID-19) pandemic has raised the need for mitigation efforts that could reduce the peak of the epidemic^1,2^. To fulfill this need, theoretical modelling can play a crucial role. Traditional epidemiology models that assume universal or constant infection parameters are not sufficient to address case specific strategies like contact tracing. Therefore, we have developed an agent-based epidemiological model which takes into account that disease transmission happens in distinct arenas of social life that each play a different role under lockdown: The family, the workplace, our social circles, and the public sphere. This subdivision becomes especially important when discussing such efforts as contact tracing. Using an estimated weight of social contacts within each of these four spheres^3^ we discuss the effect of various mitigation strategies.

At the time of writing, both classical mean field models^4,5^ and agent-based models^2,6,7^ of the COVID-19 epidemic have already been made. The models often assume contact rates and disease transmission to be stratified by age^3,8,9^. In our model, we focus on social and work networks in the spread of the epidemic. This will directly allow us to test the effectiveness of localized quarantine measures. In addition we allow a fraction of the contacts to be non-specific, representing random meetings.

Within families, several age groups may live together. At the same time, disease transmission within the family is probably the variable that is the most difficult to change through social distancing. Furthermore, there is doubt as to what extent children carry and transmit the disease^10^. By ignoring age as a factor our agent-based model implicitly weights children on equal footing with anyone else, and our model is not designed to address scenarios where one specifically targets older people.

Analysing what role each area of social life plays also allows us to separately treat social life, and since this plays a smaller economic role than work, it may be reduced with a smaller toll on society. Furthermore, if widespread testing and contact tracing is implemented, a compartmentalisation like the one we are assuming here will help in assessing which people should be quarantined and how many will be affected at any one time.

In the following, we will investigate two closely related questions. First, how a lockdown is most effectively implemented, and second, how society is subsequently reopened safely, and yet as fast as possible. To answer the first, we must examine the relative effects of reducing the amount of contacts in the workplace, in public spaces, and in closely connected groups of friends. For the second question, we will look for viable strategies for mitigation that do not require a total lockdown. Here, we will focus on the testing efficiency and contact tracing^11^. Our results will hopefully be helpful in informing future containment and mitigation efforts.

## Methods

Our proposed model divides social life into family life which accounts for 40 % of all social interactions, work life accounting for 30 %, social life in fixed friend groups accounting for 15 %, and public life which accounts for another 15 %^3^. Interactions in public are taken to be completely random and not dependent on factors such as geography, density or graph theoretical quantities. Within families, workplaces, and friend groups, everyone is assumed to know everyone. Each agent is assigned one family and workplace, as well as two groups of friends. Workplaces on average contain ten people, whereas each friend group on average contains five. In the simulation runs presented here, we use a population of *N* = 5000 agents. Increasing the number of agents changes the outcome very little, except for minimising stochastic noise. We also do not allow migration in or out of the system.

We use a discrete-time stochastic algorithm. At each time-step (0.5 days), each person has one interaction with some other person. A “die roll” decides whether the person will interact with family, friends, work, or the public. The respective odds are the above mentioned percentages 40:30:15:15. If the public is chosen, an entirely random person is selected, otherwise a person is drawn from a predefined group (family etc.). For each interaction, an infectious person has a fixed probability of passing on the disease to the person they interact with.

The family size distribution of is based on the distribution of Danish households^12^. The average number of people per household is approximately 2, and large households of more than 4 people have been ignored, as they account for less than 10 % of the population. We believe that in a country where family sizes are larger and there are fewer singles, the family would be more important to the spread of disease. We test the effect of larger families in the supplement and find that it does not chance our overall conclusions.

We simulate the progression of disease using an SEIR model with four exposed states, *E* = *E*_1_ + *E*_2_ + *E*_3_ + *E*_4_, each lasting on average 1.25 days, corresponding to a mean incubation period of 5 days. The exposed states are presymptomatic, meaning that people will not get tested in the incubation period. We let stages *E*_3,4_ be as infectious as the *I*-stage, as data suggest that a substantial fraction of COVID-19 transmission happens before the onset of symptoms.^13^ Multiple exposed states are included in order to get a naturalistic distribution of incubation periods. Li *et al*.^10^ report that the mean incubation period is approximately five days and the reported distribution is fitted well by the gamma distribution we obtain from our four *E*-stages.

A further problem is the duration of the infectious period (I). Viral shedding has been observed to last up to eight days in moderate illness^14^. On the other hand, according to Linton *et al*., the median time from onset to hospitalisation is three days^15^. A bedridden patient (even if not hospitalised) is likely to transmit the disease less. To fit the observed mean serial intervals of 4.6 days of Nishiura *et al*.^13^ we model the infectious period as a single state with an average duration of three days. In addition, the infectious, presymptomatic period lasts on average 2.5 days. In comparison ref.^16^ uses a serial interval distribution with mean of 6.5 days. Other authors have suggested a longer serial interval^10^ with presymptomatic infections.

Finally, the transmission rate for the disease is estimated from an observed rate of increase of 23 % per day in fatalities in the USA. This also fits the observation of a growth rate of ICU admissions of about 22.5 % per day in Italy^17^. With our parameters this is reproduced by a basic reproduction number *R*_0_ *∼*3 (as we allow transmission in both directions when selecting two people, this is simulated by a rate for transmission of 0.2 per encounter). Li *et al*.^10^ estimate *R*_0_ at 2.2 based on a growth rate of 10 % per day in confirmed COVID-19 cases in Wuhan prior to Jan. 4.

Having calibrated the model in this way, we want to explore mitigation strategies for the corona epidemic. Specifically, we will investigate the relative importance of the areas of social life, and the extent that reducing workplace size reduces disease spread. Moreover, we will examine the possible gain and cost by simple contact tracing and light quarantine practices.

## Results: Mitigation strategies

To illustrate the relative importance of the workplace and public life, we consider the four scenarios in Fig. 2(a). In the first scenario, nothing is done. In the second, contacts within the workplace are reduced by 75 %, while in the third, contacts with friends and the public are reduced. Finally, we compare these with similar scenarios, but where good hygiene or keeping a distance reduces the probability of infection from all types of encounters by half.

**Figure 1.**
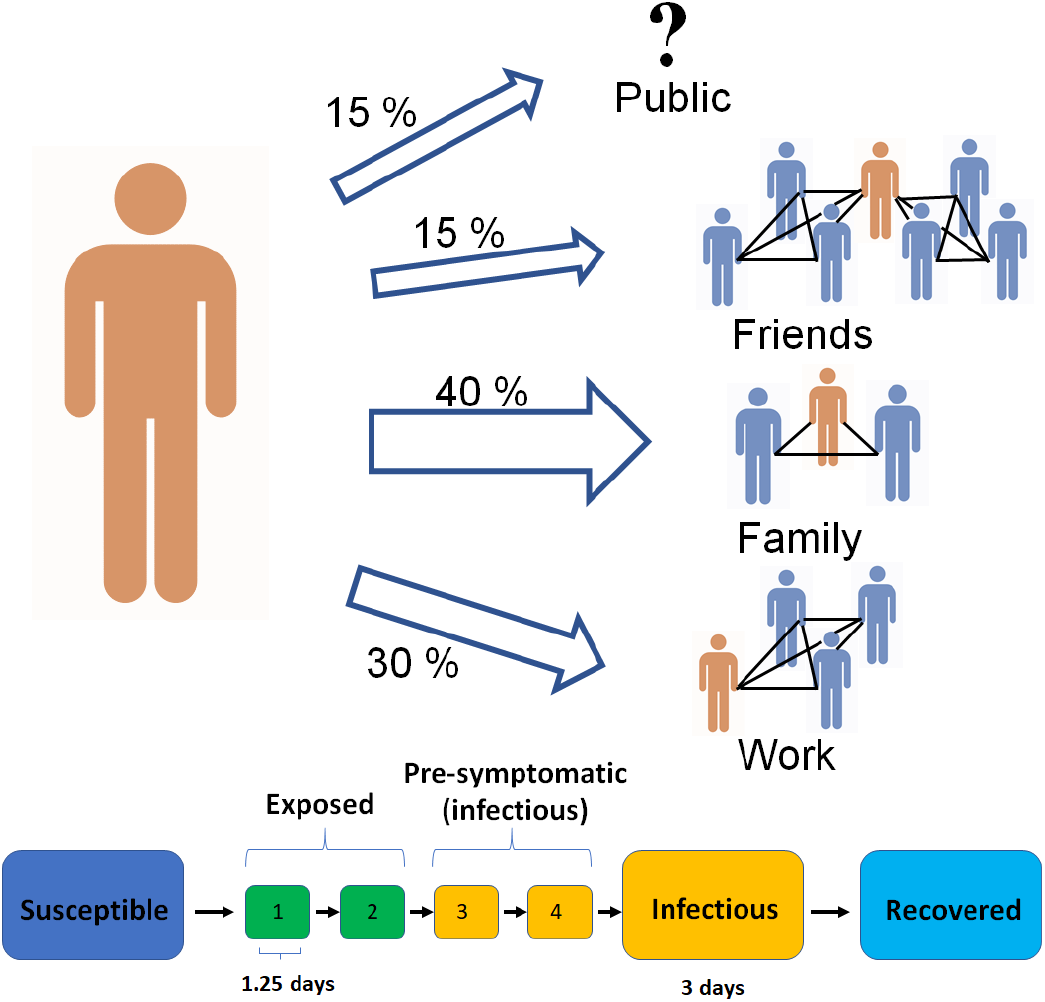
A diagram of the model structure. Each agent has a network consisting of a family, a workplace and two groups of friends. The family accounts for 40 % of interactions. Work accounts for 30 % and socialisation with friends accounts for a further 15 %. The links in each of these 3 groups are fixed throughout the simulation. Finally, 15 % of interactions happen “in public”, which we implement as an interaction with a randomly chosen other agent. Everyone in the work and friend sub-graphs are assumed to be connected to each other. Below the graph, the underlying mechanisms of the disease are shown. We divide the exposed state into four in order to get a more naturalistic gamma distribution of incubation periods. The two last exposed states are infectious, but asymptomatic, meaning that individuals will not get tested. This is to include presymptomatic infection. In our simulation we set the family groups to an average of 2 people, and the work network to 10 completely interconnected people. The friend network consist of 2 groups with 5 in each.

**Figure 2.**
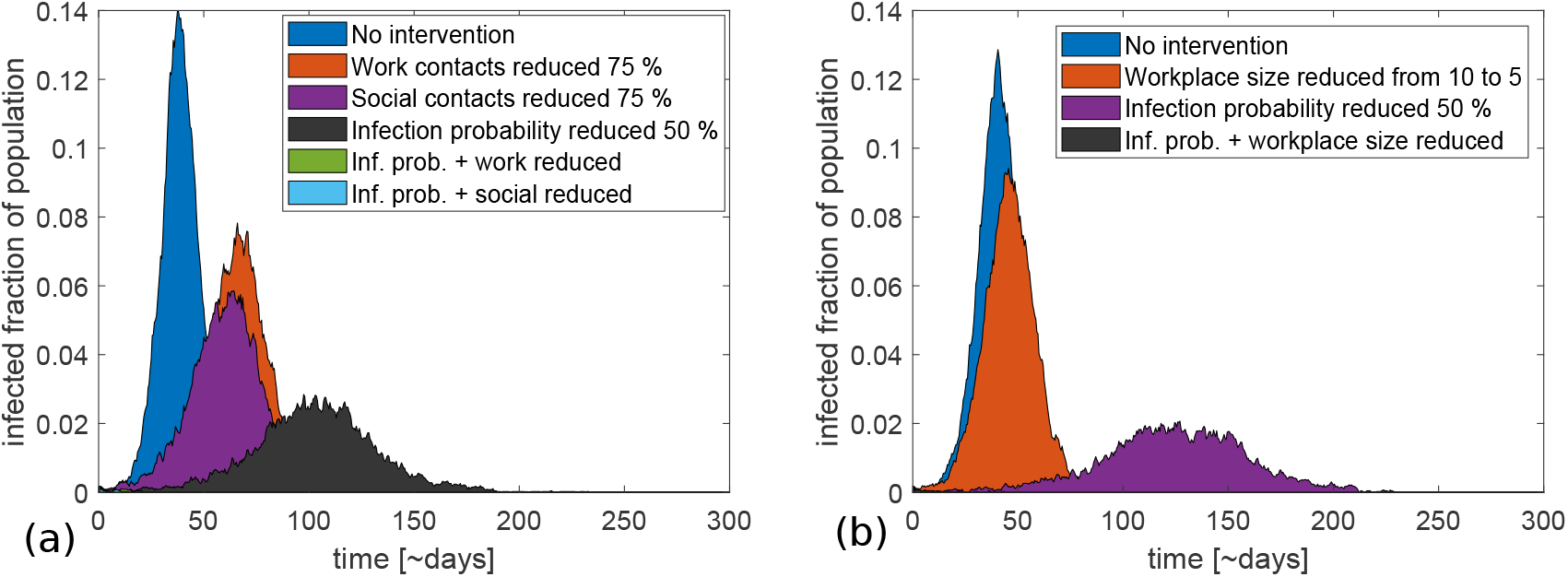
Comparison of various strategies with and without a reduction in transmission probability per encounter. (a) Reducing contacts in different social contexts by 75 % through a lockdown. It can be seen that work and social contacts play roughly the same part in disease transmission. A reduction of infection probability makes the strategies relatively more effective. Reducing work contacts, for example, reduces the peak height by roughly 40 % (relative to no intervention) if infection probability is high. If the infection probability is lowered, the strategy completely eliminates the epidemic. The apparently missing lines in the figure are due to the epidemic dying out completely when a lockdown is combined with hygienic measures. (b) A similar comparison of the effects of reducing workplace sizes by half. This strategy is also relatively more effective if infection probability is reduced. The strategy reduces peak height by 75 % at a lowered infection probability versus only 35 % if infection probability remains high.

In the figure, we see that the effects of reducing workplace and social contacts are roughly of the same magnitude. This reflects the assignment of 30 % weight to each of these contact types. The slightly larger effect on social contacts reflect our assumption that these connections are less clustered than the workplace network. The two latter graphs show the scenarios where we both reduce infection probability within one group by 75 % and overall infection probability by 50 %. They show that an effective lockdown requires both restrictions of the time spent in the workplace and in the public sphere, and measures that reduce infection probability by increased hygiene and physical distancing.

The above results provide one useful piece of information. If the effect of workplace and social contacts are of the same order, it is of little importance which one is restricted. Ideally, both will be restricted for a period of time. However, when restrictions need to be lifted, authorities will primarily be able to control the workplace, whereas the social sphere relies on local social behaviour. Obviously it is economically more sustainable to lift the one with the largest societal consequences first, by allowing people to return to work while encouraging keeping social gatherings at a minimum.

If restrictions are lifted before herd-immunity has grown to substantial levels, the epidemic will re-ignite. Therefore, we now examine what can be done to minimise spread in the reopened workplaces.

One possible strategy is to reduce the number of people allowed at any one time in each workplace. In Fig. 2 (b), we compare an epidemic scenario where the average number of employees per workplace is 10 with an epidemic where this number is reduced to 5. We further assume that the number of contacts per coworker remains the same, meaning that the number of contacts per person drops when workplace size is reduced.

It can be seen that fragmentation of physical spaces at workplaces could have a significant effect on the peak number of infected. In a situation with a risk of straining the healthcare system, this could be part of a mitigation strategy. Once again, the strategy becomes relatively more effective if the infection probability per encounter is also reduced. Relative to the cases with no workplace size reduction, making workplaces smaller leads to a greater relative reduction in peak size if infection probability is lower (12 % versus 28 % relative reduction when when overall infection rates are halved by other means).

A more local strategy that can be employed when reopening society is widespread testing and contact tracing. As mentioned above, Hellewell *et al*.^18^ have suggested that this can be effective in containing COVID-19 outbreaks provided high efficiency in detecting infected individuals. Contact tracing has previously been modeled in relation to other epidemics^19^, and used successfully against smallpox^20^ and SARS^21^.

One obstacle to the widespread implementation of this strategy is difficulty of tracing contacts. Therefore, we will here implement a crude form of contact tracing where we 1) close the workplaces of people who are tested positive for the disease, isolate their regular social contacts for a limited period, and 3) keep symptomatic individuals in quarantine until they recover. We will see that such a 1 step tracing and quarantine strategy (1STQ) can give a sizeable reduction in disease spread while costing fewer lost workdays than overall lockdown. Our simulations include the limitations imposed by not being able to trace the estimated 15% of infections from random public transmissions. Thus the strategy does not require sophisticated contact tracing but could be implemented based on infected people being able to recollect their recent physical encounters with friends. It should be noted that we here quarantine persons in their own households, thereby making our contact tracing strategy easier to implement in practice. In particular, family members of a quarantined person are still free to interact outside their home if they are not themselves tested positive. The drawback of such light quarantine practices is that infected persons in quarantine may still transmit the infection to their families.

Fig. 3 examines how increased detection efficiency systematically improves our ability to reduce the peak disease burden. This would then be a more cost efficient way to mitigate the pandemic than a complete lockdown where each person would lose several man-months. Even detecting as little as 5 % of COVID-19 infected per day (which with an average symptomatic disease duration of 3 days corresponds to finding approximately 15 % of the infected) can potentially reduce the peak number of cases by 50 %. If 10 % efficiency is possible, corresponding to detecting about a third of infectious cases, then peak height could be reduced by a factor 3, and with less than two weeks in quarantine per person during the entire epidemic. This is illustrated in Fig. 3(a) where peak height is reduced from 0.13 to 0.04 at 10 % testing efficiency.

**Figure 3.**
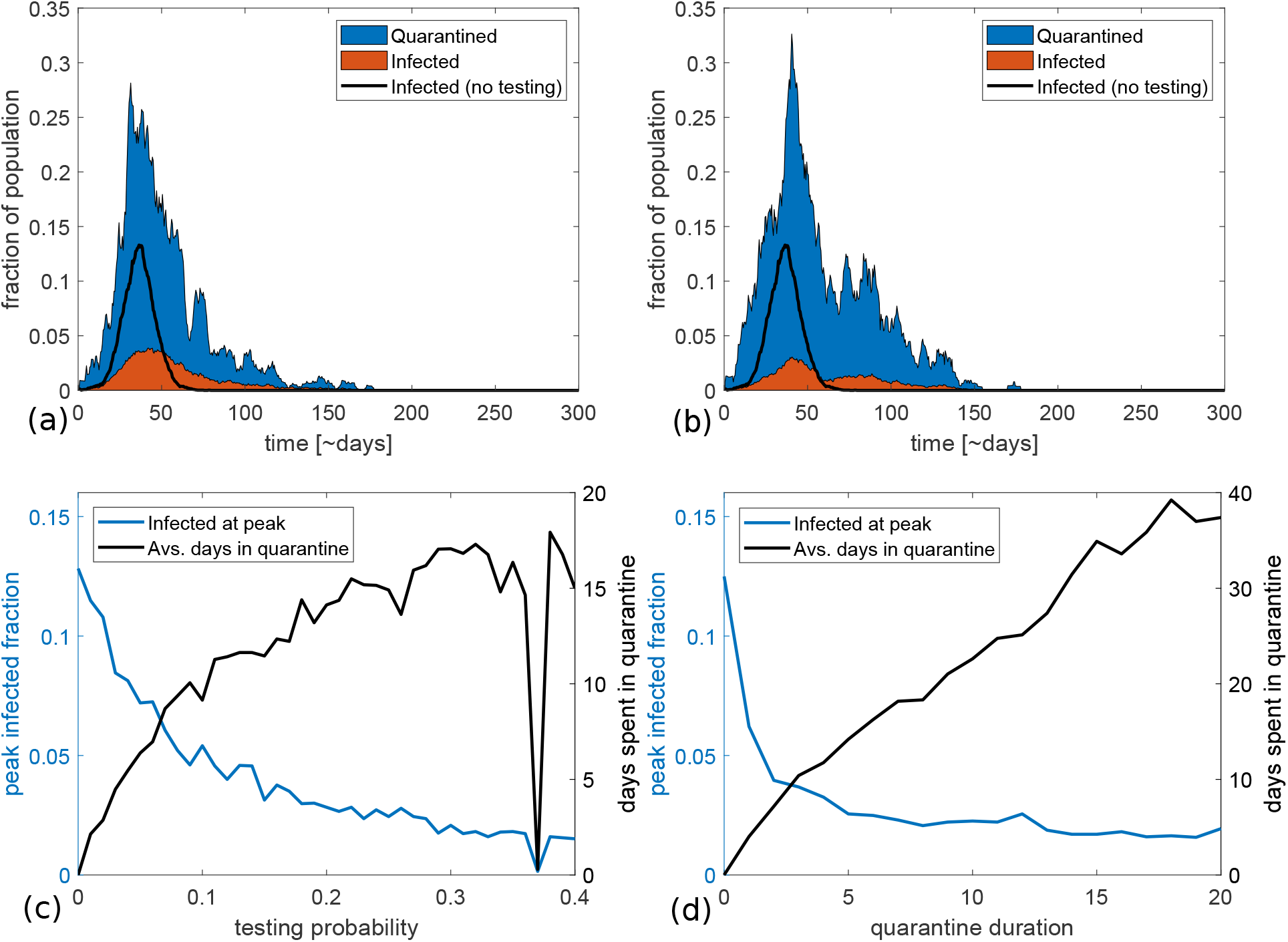
The effect of quarantine duration and testing probability. (a) and (b) show examples of epidemic trajectories for a quarantine length of 5 days and a daily testing probability of 10 and 20 % respectively. The blue section of the curve shows the fraction of people who are in quarantine but healthy or presymptomatic, while the orange section shows the fraction who are ill. (c) and (d) show the peak fraction of population infected (left y-axis) and average time spent in quarantine (right y-axis) as a function of testing chance and quarantine length. The number of days in quarantine was calculated using our standard group sizes which connect each person to approximately 20 others. The average quarantine time scales proportionally with this assumed connectivity. (c) With a quarantine length of 5 days, it is possible to reduce the peak number of infected by eight percentage points, corresponding to a 60 % drop, if the probability of infected people being tested is only 10 % per day of illness. However, the price of this is that each person is on average quarantined twice during the epidemic. If testing is more widespread, the epidemic peak can be reduced, until it finally becomes unstable at a testing probability of around 40 % per day. If it is possible to completely eliminate the epidemic, the number of days spent in quarantine is similarly reduced. (d) Epidemic peak and time spent in quarantine as a function of quarantine length for a testing probability of 20 % per day. The average time spent in quarantine increases linearly with the length of quarantine. On the contrary, the effect of quarantine on the peak height appears to stagnate at approximately five days.

The main cost of the quarantine option is the quarantine time. Figure 3(d) examines the efficiency versus cost of as a function of quarantine length. It can be seen that there is little gain in extending the quarantine period beyond the 5-day duration of the incubation period. For this reason we opted for 5 days in quarantine in panel (a,b). As a consequence, an average person will stay around 12 days in quarantine during the course of the epidemics with a testing efficiency of 10 %. This time can be reduced if people can convinced of smaller work environments and fewer physical contact per week. Fragmentation of our networks into smaller groups will reduce both quarantine overhead and the direct transmission of the disease (Fig. 2(b), orange curve).

A prolonged lockdown will hugely disrupt society, and it is questionable whether a complete eradication of the virus is possible anyway. Therefore most governments have aimed at softening the epidemic curve, with varying degree of success. The here explored one step contact tracing with testing and quarantine is a means to this end, and would work most efficiently in combination with other means to reduce *R*_0_.

Finally, we investigate whether an aggressive testing and contact tracing strategy could work if implemented at a late stage in an epidemic. This could be relevant if for example the strategy is part of an effort to reopen society after a period of lockdown.

In fig. 4, we show two possible scenarios where testing and contact tracing is implemented after a 30 day lockdown with a 75 % reduction of work and social sphere. The lockdown is initiated when 1 % of the population is infected. In (a) we subsequently test and quarantine the infected and their contacts for 5 days, while in (b) the required quarantine is set to 10 days. We assume a testing efficiency of 20 % chance of detection for each day a person is symptomatic. The progression of the epidemic without testing is marked by a black graph for comparison.

**Figure 4.**
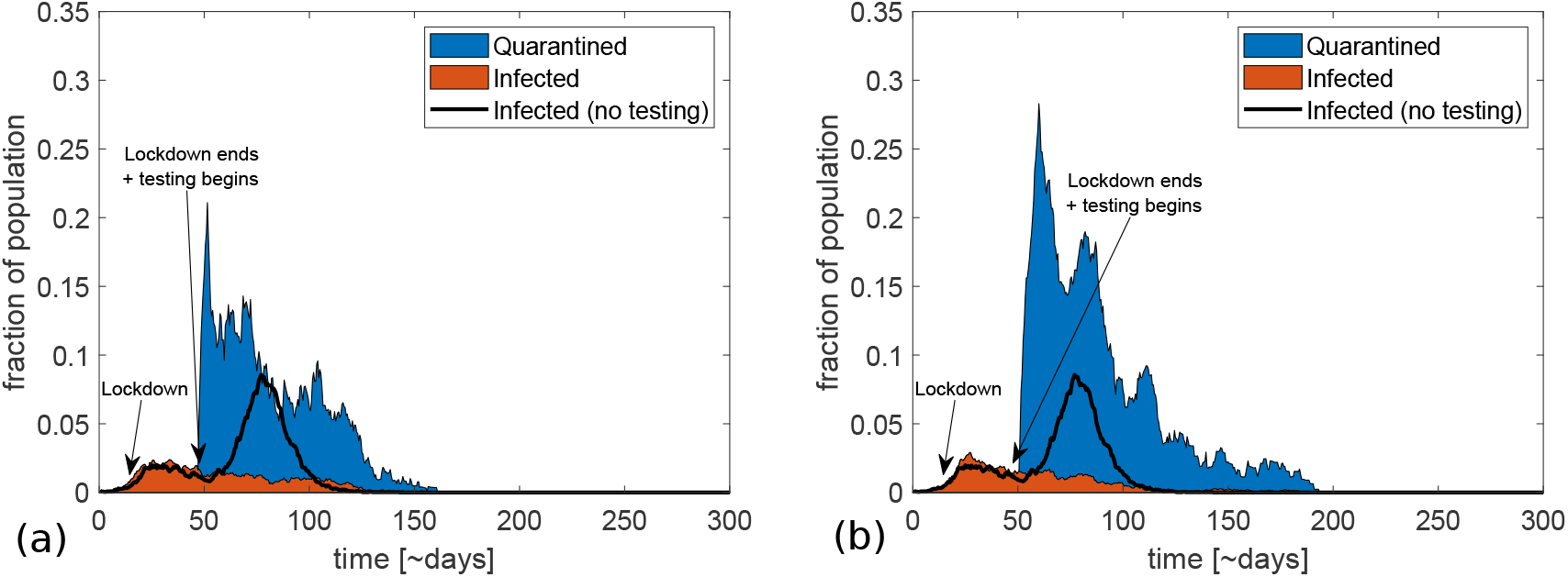
Various trajectories of the epidemic when combining a lockdown and a late-onset 1STQ strategy. (a) shows a possible course of an epidemic where restrictions in public and work life (by 75 %) are implemented when 1 % are infected and lifted after 30 days, being replaced by a testing and tracing regime with a testing probability of 20 % per day and quarantine duration of 5 days. The black line shows the fraction of infected if no testing is implemented. We see that this level of testing and quarantine is sufficient to prevent a resurgence of the epidemic. (b) is similar, but here quarantine lasts 10 days. This is about as effective as (a), but costs a lot more in terms of number of people in quarantine.

From the figure one sees that the replacement strategy of even relatively short quarantines also works with a late onset. At a realistic detection probability, it prevents a resurgence of the epidemic. Nonetheless, it is quite costly initially, with a very high peak in number of quarantined people. Most importantly, the effect does not increase with a longer quarantine period, but the cost is substantially larger.

## Discussion

Pandemics such as the one caused by COVID-19 can pose an existential threat to our social and economic life. The disease in itself is serious, and leaves specific epidemic signatures and characteristics that make traditional contact tracing difficult. In particular it is highly infectious, can sometimes be transmitted already 2 days after exposure, and a large fraction of transmission happens before the onset of symptoms. As such it is difficult to contain without a system-wide lockdown of society. Nonetheless, a successful containment in South Korea used contact tracing. This motivated us to explore a one-step contact tracing/quarantine strategy (1STQ).

Using reasonable COVID-19 infection parameters we find that the 1STQ strategy can contribute to epidemic mitigation, in the sense that it can reduce the peak number of infected individuals by about a factor 3 with realistic testing rates. This was illustrated systematically in Fig. 3. The main cost was people in self-quarantine and not contributing to the workforce. In comparison one has to consider that a society-wide lockdown with similar reduction in peak height would have to last for about 100 days (see Fig. 2). Thus, the lockdown would require of order 100 days of quarantine (or at least extensive social distancing) per person, whereas testing and isolation only requires on average around 12 days per person with a 5-day quarantine. Importantly, these numbers can be reduced if people are able to lower their number of contacts.

A noticeable objection to the 1STQ strategy is the fraction of cases with so weak symptoms that people do not contact health authorities. The effect of such limitations is in our model parameterized through the detection probability. From Fig. 3(c) one sees that when the detection probability goes below 3 % (a rate of 1 % per day) the peak reduction of the 1STQ strategy becomes only of the order 1 percentage point. It should also be noted that, since we rely on symptoms to determine who stays in quarantine, and people in the infectious/symptomatic stage are assumed to always stay in quarantine, we implicitly assume that all infected persons develop at least some symptoms at some point. This may be a break from reality.

The increasing availability of tests may also change the perspectives of the 1STQ strategy. With widely available rapid tests, it will be possible to test everyone regularly, and to test all quarantined persons before they leave quarantine. Supplementary figure S4 deals with the results of such a testing strategy. To put this into perspective, the drawbacks of widespread, but slow testing is examined in the supplementary figure S5.

One interesting point which we have not examined here, is that real-world social networks are heterogeneous, with a large variance in number of contacts. It may be expected, for example, that workers in customer-facing positions in shops will have a high risk of catching the disease and passing it on. The effects of this heterogeneity is examined more closely in ref.^22^. Here, it is concluded that heterogeneity in the number of contacts enhances the effect of contact tracing, since persons with many contacts are both more likely to pass on the disease and more likely to be quarantined.

In ref.^18^ the authors suggest a 1STQ strategy similar to the one we here model. The main points of the present analysis is the focus on mitigating instead of eradicating the epidemic, our suggestion of a shorter quarantine length, and the implementation of quarantine together with other members of the household instead of total isolation. Our stochastic approach also allows for local failures due to the limited duration of quarantine (people may not yet be symptomatic when exiting quarantine) and the non-traceable public contacts (set to 15%).

Finally, one noticeable finding is that contact tracing and reduction of contacts per person is still feasible even at a later stage of the epidemic. As can be seen in Fig. 4, a lockdown and a subsequent reopening with testing and contact tracing is highly effective in controlling the epidemic. Our study thus shows that lockdowns have an important role to play in epidemic mitigation, but that they can be replaced by a 1STQ strategy once the epidemic is under control.

The COVID-19 pandemic has set both governments, health professionals, and epidemiologists in a situation that is more stressful and more rapidly evolving than anything in recent years. Due to the uncertainties caused by a situation in flux, it is difficult to predict anything definite about what works and what does not. The empirical observation that lockdowns worked in both China, and in a milder form in Denmark shows that our assumption of a 75 % reduction in specific infection rates under lockdown is realistic. Our main result is that some of these restrictions can be replaced by testing, 1 step contact tracing and short periods of quarantine. This is far cheaper than total lockdowns. Perhaps most importantly, these measures work best in combination. As is highly relevant to the current epidemic stage of COVID-19, we pinpoint that 1STQ can be successfully implemented also at a late stage of the epidemic where testing may become massively available.

## Data Availability

The algorithm used to carry out the simulations described in the manuscript is available on figshare (https://doi.org/10.6084/m9.figshare.12206735.v4)

## Acknowledgements

We thank Gorm Gruner, Bjarke Frost Nielsen, Andreas Roepstorff and Lone Simonsen for enlightening discussions. This project has received funding from the European Research Council (ERC) under the European Union’s Horizon 2020 research and innovation program under grant agreement No. 740704.

## Author contributions

AE and KS both participated in devising the model. Code was written and plots produced by AE. The functionality of the code was checked by comparison with an alternative algorithm written by KS. AE and KS wrote and edited the manuscript.

## Competing interests statement

The authors declare no competing interests.

## Data availability

Plots of alternative variants of our model (including larger family sizes and alternative quarantine strategies) can be found in the supplementary material. The code used to produce the plots shown in this article is available on Figshare under the URL https://doi.org/10.6084/m9.figshare.12206735.v4.

## Supplementary figures for the article

### The effects of family size

To examine the effect of the family size distribution on our conclusions, we have repeated the figures of the main article with twice as large families. That is to say, single households now contain two people, previously two-person households contain four etc. We see that while this somewhat affects the shape of the epidemic curve, it does not significantly influence any of our conclusions.

**Figure S1.**
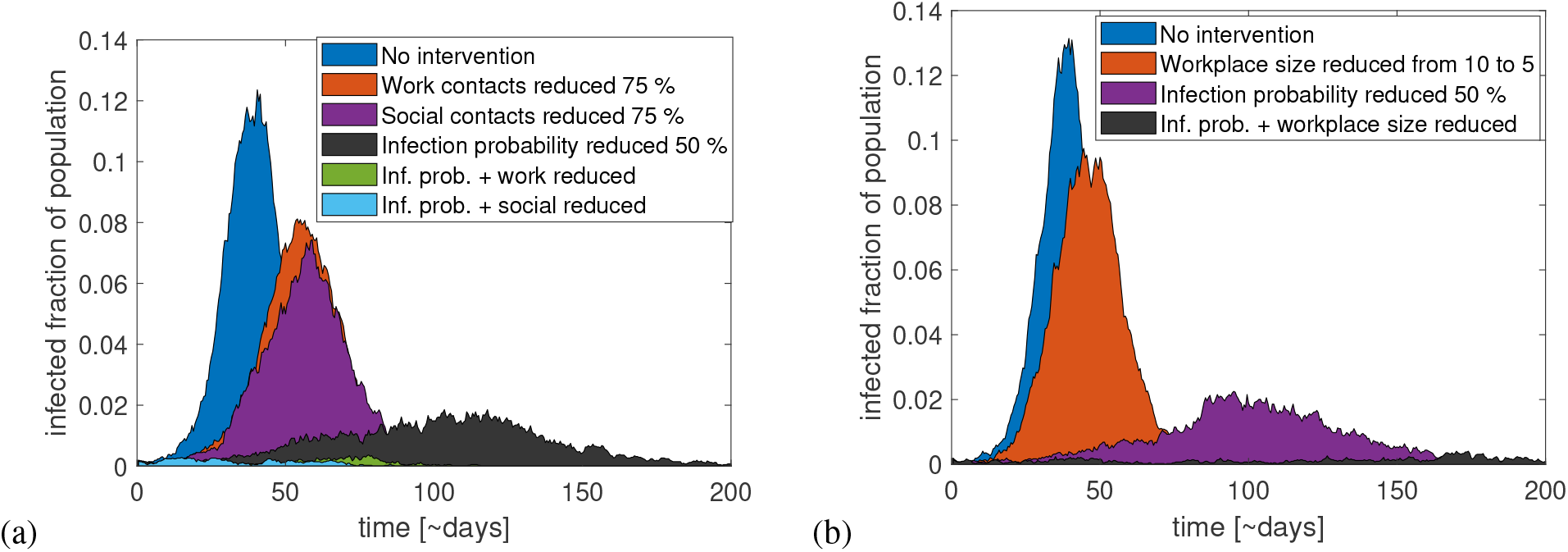
(a) A comparison of various containment strategies that include a lockdown reducing social or work contacts by 75 %. We see that if we combine these measures with improved hygiene or social distancing, reducing the transmission risk per encounter by half, this is sufficient to stop the epidemic completely. This is the same conclusion as in the main article. (b) The effect of reducing workplace sizes by half, from 10 to 5 people per workplace on average. As in the main article, this has a significant effect on the epidemic, and in this version of the model, it is also enough to fully mitigate it if combined with hygienic measures that halve the infection probability.

**Figure S2.**
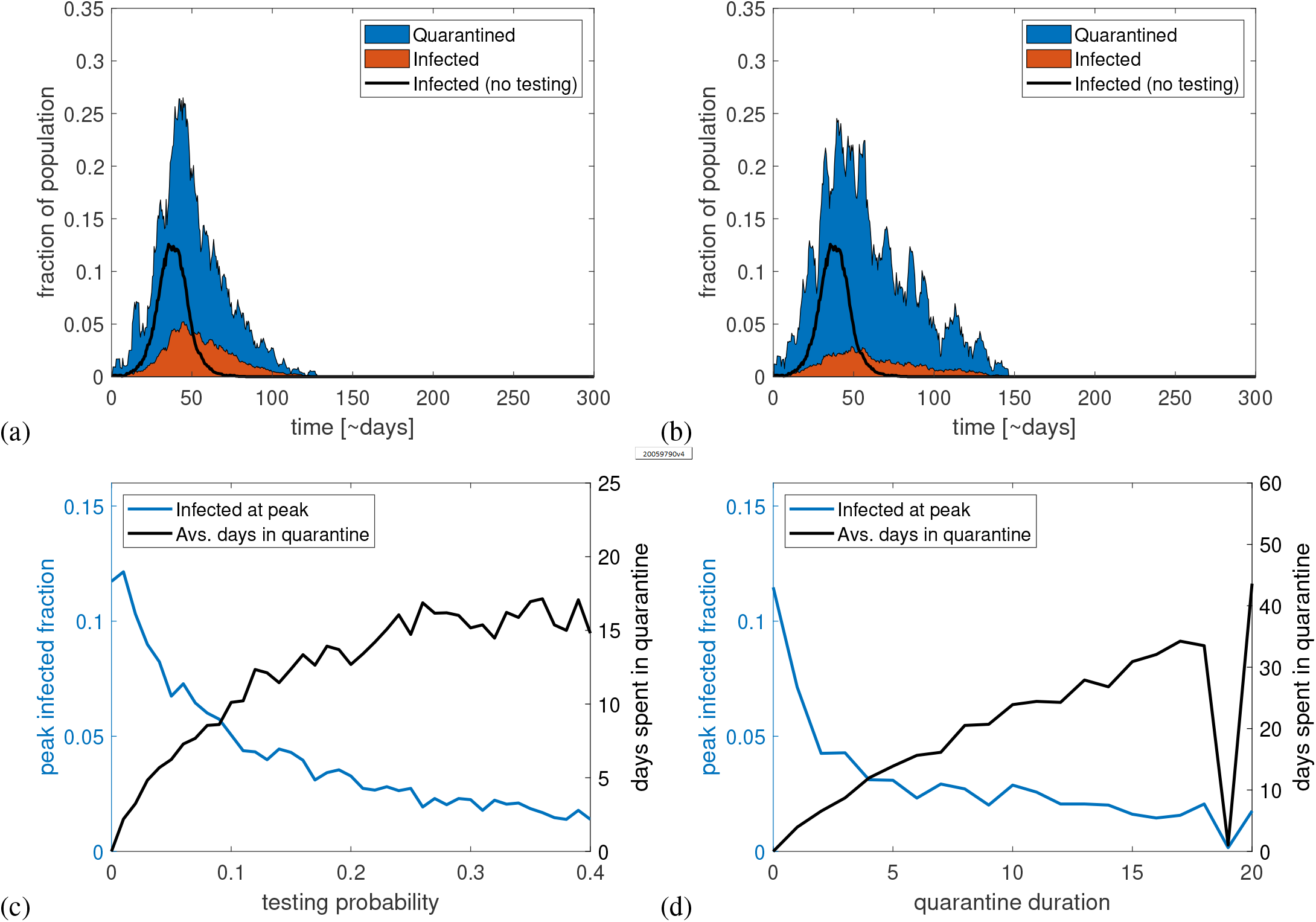
(a) and (b) show epidemic trajectories and fraction of people in quarantine for testing probabilities of 10 and 20 % per day respectively. The black line shows the epidemic trajectory in the absence of testing and contact tracing. (c) shows the infected fraction of the population at the peak of the epidemic (left axis) and the average number of days each person spends in quarantine during the epidemic as a function of daily testing probability while symptomatic. (d) shows the same variable but as a function of quarantine duration. It can be seen here as well that little is gained from quarantine beyond five days. When quarantine lasts longer than about 18 days, the epidemic becomes unstable, sometimes dying out.

**Figure S3.**
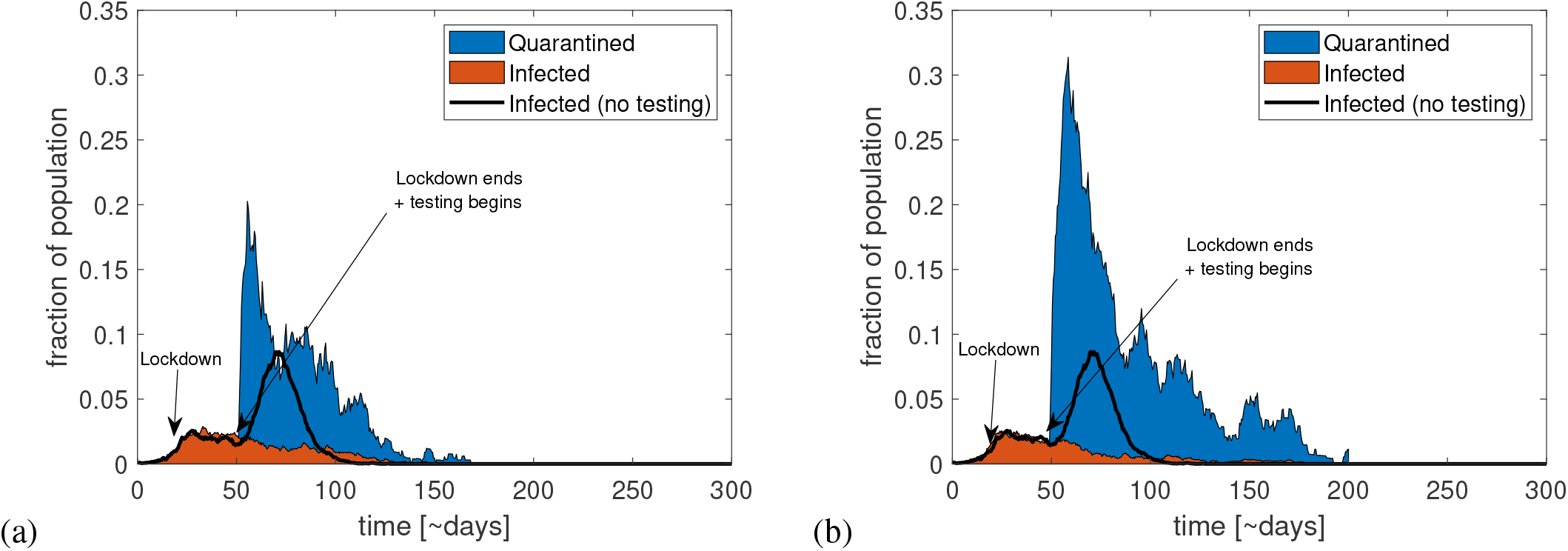
Two examples of epidemic trajectories when combining a lockdown with a 1STQ strategy. When 1 % of the population is infected, a lockdown is implemented, reducing social and work contacts by 75 %. It is lifted after 30 days and replaced by a testing and contact tracing strategy with a daily testing probability of 20 % for symptomatic individuals. The duration of the quarantine in panel (a) is five days and in (b) it is ten days. We see that the longer quarantine does not change the effect on the epidemic much, but it does increase the number of people in quarantine.

### Weekly tests

In this section, we let all agents get tested regularly at a one-week interval in addition to a 20 % daily testing probability for symptomatic individuals. This strategy will become increasingly feasible with increasing availability of rapid tests. It can be seen that weekly tests are sufficient to contain the epidemic if contacts of the infected are quarantined (fig. S4(a)). If only the infected themselves are quarantined, it is still enough to mitigate the epidemic as seen in panel (b), giving us a situation where no healthy persons are unnecessarily quarantined. With as widespread testing as this, it is assumed that everyone is tested before leaving quarantine, meaning that no presymptomatic individuals are let out of quarantine.

**Figure S4.**
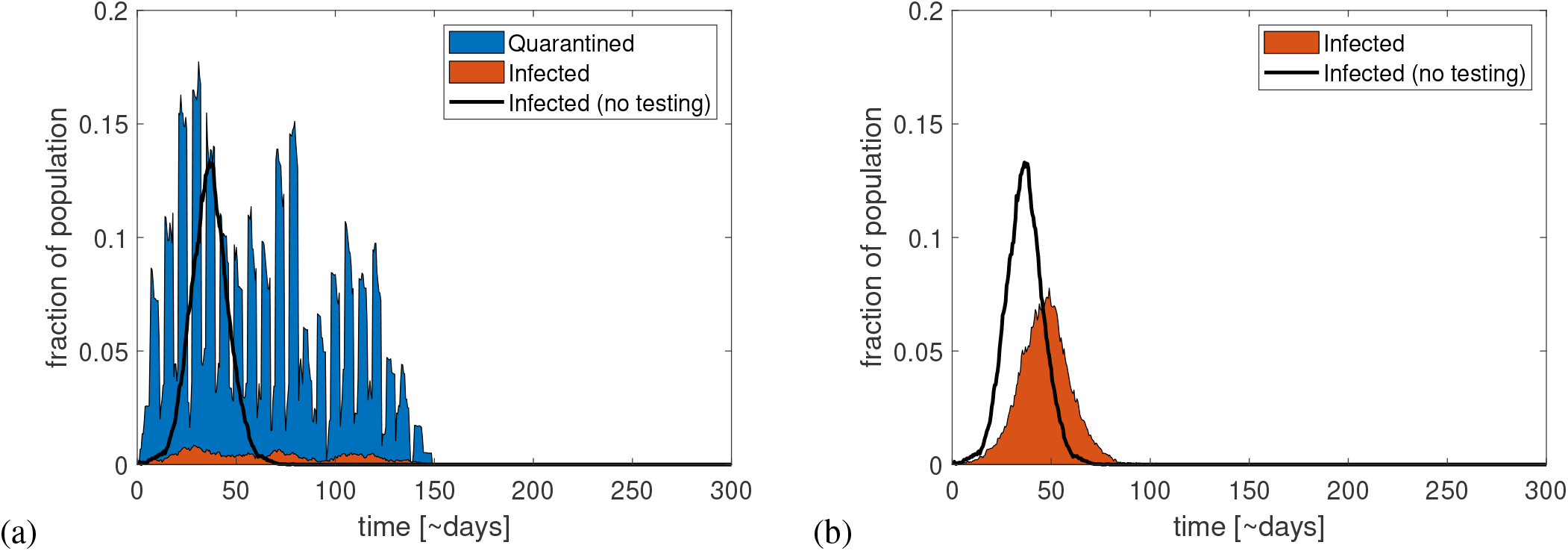
(a) Infected and quarantined fraction of the population if everyone, in addition to tests of the symptomatic, take one weekly test, thus also discovering any presymptomatic cases. This is enough to keep the epidemic in check. In (b) the same strategy is followed, but only the infected themselves are quarantined. This still significantly reduces the epidemic peak. The unmitigated epidemic trajectory is shown by the black curves.

### Delayed test results

In the following, we examine the effect of a delay in obtaining test results. We assume that people who are tested have to wait for a number of days before getting the result, thus delaying contact tracing and quarantine. We find that the effect of contact tracing only subsides slowly with increasing test delay, as seen in fig. S5. This we believe is due to the effect of the large number of people in quarantine at the peak of the epidemic. Even though the contact tracing efforts are less effective when test results are delayed, having a large fraction of the population in quarantine works as a primitive lockdown, lowering the epidemic peak by “brute force”.

**Figure S5.**
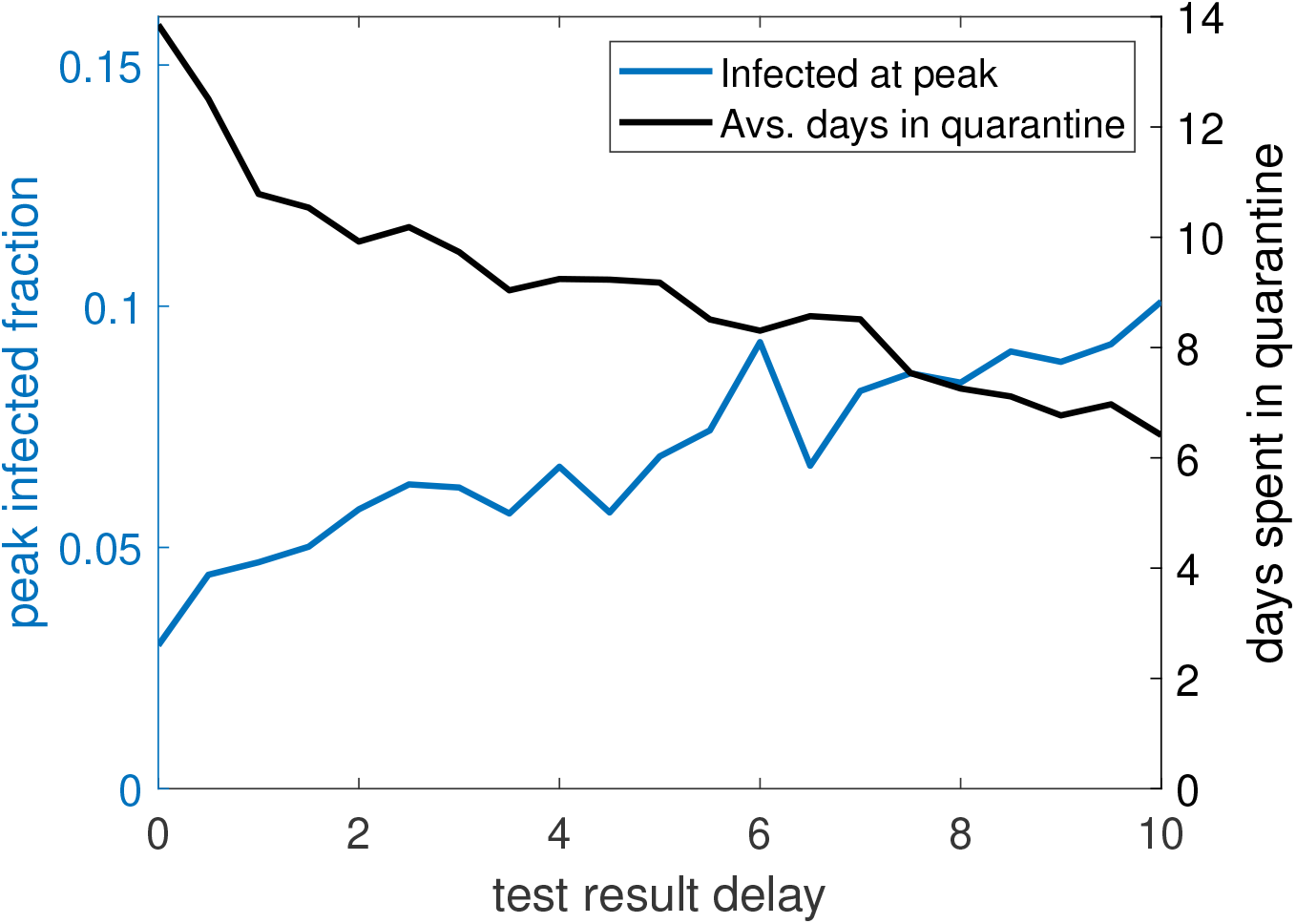
This figure shows the effect of delayed test results on the peak fraction of infected and the average number of days spent in quarantine during the epidemic. We see that the peak fraction of infected grows linearly with the delay, until the effect is nearly gone after ten days. At the same time, the amount of time spent in quarantine decreases. Part of the mitigation at long delays is expected to stem from the fact that a significant portion of the population is quarantined, and therefore less infectious, at all times. The testing probability is here set to 20 % per day of symptomatic illness and the quarantine length is set to five days. The testing delay is assumed to not affect people’s ability to leave quarantine.

## Notes

### Competing Interest Statement

The authors have declared no competing interest.

### Summary of Updates

Updated manuscript and title to match with the latest revision currently under review. Added new supplement examining the effect of family size, testing at regular intervals, and delayed test results.

